# The protection gap under a social health protection initiative in the COVID-19 pandemic: A case study from Khyber Pakhtunkhwa, Pakistan

**DOI:** 10.1101/2022.11.29.22282883

**Authors:** Sheraz Ahmad Khan, Kathrin Cresswell, Aziz Sheikh

## Abstract

**Background:** Sehat Sahulat Programme (SSP) is a Social Health Protection (SHP) initiative by the Government of Khyber Pakhtunkhwa (GoKP), covering inpatient services for 100% of the province’s population. In this paper, we describe SSP’s role in GoKP’s COVID-19 response and draw inferences for similar programmes in Pakistan.

**Methodology and methods:** We conceptualised SSP as an instrumental case study and collected three complementary data sources. First, we studied GoKP’s official documents to understand SSP’s benefits package. Then we undertook in-depth interviews and collected non-participant observations at the SSP policy and implementation levels. We recruited participants through direct (verbal and email) and indirect (invitation posters) methods.

Use of maximum variation sampling enabled us to understand contrasting views from various stakeholders on SSP’s policy dimensions (i.e., coverage and financing), tensions between the policy directions (i.e., whether or not to cover COVID-19) and how policy decisions were made and implemented. We collected data from March 2021 to December 2021. Thematic analysis was conducted with the help of Nvivo12.

**Findings:** Throughout 2020, SSP did not cover COVID-19 treatment. The insurer and GoKP officials considered the pandemic a standard exclusion to insurance coverage. One SSP official said: “COVID-19 is not covered and not relevant to us”. GoKP had stopped non-emergency services at all hospitals. When routine services restarted, the insurer did not cover COVID-19 screening tests, which were mandatory prior to hospital admission.

In 2021, GoKP engaged 10 private SSP hospitals for COVID-19 treatment. The SSP Reserve Fund, rather than insurance pooled money, was used. The Reserve Fund was originally meant to cover high-cost organ transplants. In 2021, SSP had 1,002 COVID-19-related admissions, which represented 0.2% of all hospital admissions (N=544,841).

An advocacy group representative called the COVID-19 care under SSP “too little too late”. In contrast, SSP officials suggested their insurance database and funds flow mechanism could help GoKP in future health emergencies.

**Conclusion:** The commercially focused interpretation of SHP arrangements led to a protection gap in the context of COVID-19. SSP and similar programmes in other provinces of Pakistan should emphasise the notion of protection and not let commercial interests lead to protection gaps.

## Introduction

Target 3.D of the Sustainable Development Goals (SDGs) embodied a commitment from all countries to build the capacities for early warning, risk reduction and management of national and global health risks. The International Health Regulations (IHR) and emergency response capacities were selected as indicators to measure progress on Target 3.D of the SDGs.(1) These indicators were measured through the World Health Organization’s (WHO) Joint External Evaluation of Preparedness (JEEP) of IHR and emergency response capacities.(2)

WHO conducted JEEP of Pakistan’s IHR and emergency response capacities in 2016.(2) The COVID-19 pandemic arrived in Pakistan, while the JEEP recommendations were not fully implemented.(2) The first case of COVID-19 in Pakistan was confirmed in Karachi on 26 February 2020, while the first death was reported on 18 March 2020 in Mardan, a district in the northern province of Khyber Pakhtunkhwa (KP).(3,4) KP was particularly hit-hard and had the highest fatality rate of all the provinces.(3)

Many factors are likely to have contributed to the poor COVID-19 outcome in KP. For example, the 2016 JEEP highlighted that there was one biosafety lab in KP but had no biosafety officer.(3) Tertiary care hospitals had no communication with each other.(1) The biosecurity situation was unknown, and the province did not have an inventory of dangerous pathogens. Another concern for KP was the lack of plans for private-sector engagement in responding to health threats.(5)

The Government of Khyber Pakhtunkhwa (GoKP) has taken steps to enhance the private sector’s participation in health care through the Sehat Sahulat Programme (SSP).(6) SSP is a Social Health Protection (SHP) initiative providing insurance coverage to 100% of the province’s population (around 36 million).(7,8) SSP provided inpatient care to permanent residents of KP through a mix of public-private hospitals.(6)

By the end of 2021 (when our data collection completed), SSP had a network of 165 hospitals. The network had 127 (77%) private and 38 (33%) public hospitals. In 2021 alone, SSP recorded 544,157 hospitals admissions with 372,924 (68%) admissions into private and 171,233 (32%) in public hospitals.(9) These data suggest that the private sector played a significant role in serving SSP patients.

There was a research gap on whether SSP, especially its private sector providers, played a similar role in responding to the COVID-19 pandemic. This study was undertaken to bridge this evidence gap by: (i) exploring SSP’s response, especially its private hospitals’ role in responding to the COVID-19 pandemic; (ii) identifying gaps in SSP response; and (iii) exploring possible options to enhance SSP’s role in responding to future epidemics/pandemics.

This paper is part of a four-paper series, where each paper is explicitly related to different dimensions of the programme. In one paper (already published), we have contextualised SSP in the broader context of UHC in KP.(10) In two papers, we will describe our findings related to the notion of access under SSP (achievements and challenges) and the role of the German Development Bank as SSP’s policy entrepreneur.

## Methodology and methods

### Study design and ethics approval

We employed an instrumental case study design approach. We had ethics approval from the University of Edinburgh (UK) and Khyber Medical University (Pakistan). We complied with the ethics regulations. We had written informed consent from all participants and ensured participants’ autonomy, confidentiality, and right to withdraw.

### Data sources

We used three complementary data collection methods. First, we reviewed governmental programme documents (Appendix 1). Second, we undertook in-depth interviews with key stakeholders (Appendix 2), and finally, we undertook non-participant observations at SSP policy level meetings and hospital-based implementation sites (Appendix 3).

### Sampling and recruitment

We acquired the documents from the SSP head office and its official website. The included documents were either authored or commissioned by the KP, including Planning Commission Form-1 (PC1) and the contract between GoKP and State Life Insurance Corporation (SLIC). A PC1 is a detailed policy and operational document approved in advance for each government programme in Pakistan. The plans envisaged in PC1s were binding on the programmes they entailed.

We used purposive (maximum variation) sampling for conducting interviews and collecting observations. The maximum variation sampling helped us to have strategic (policy level) and operational (implementation) level views. We were able to compare and contrast views from our respondents on SSP’s policy directions during the pandemic (like coverage and financing strategies) and tensions between the policy parameters (whether or not to cover COVID-19). We were also able to understand how policy decisions were made at the strategic level (SSP head office) and how they translated (or did not translate) into action(s) at the hospitals.

Key stakeholders in our study included officials from GoKP, the insurance company, officials of the SSP network hospitals, public advocacy groups and technical experts working at international development agencies in KP. We recruited participants through direct (face-to-face or written communication) and indirect invitations (by displaying invitation posters in stakeholders’ offices). For collecting observations, our access to the meetings at the SSP head office and SSP desks in hospitals was facilitated by the SSP Director.

### Data collection

We collected data from March 2021 to December 2021. Informed, written consent was taken from all participants. We collected data as per the interview guide and the observation sheet that we had developed. The programme documents were used to follow the SSP evolution in terms of the population, services and financial coverage. Through the interviews with stakeholders, we explored the reasons behind the policy decisions (about COVID-19 coverage), their implications like out-of-pocket (OOP) expenditure and the potential ways to harness SSP’s role in responding to any future pandemics. Through the non-participant observations, we explored how SSP policy decisions were made and how they were (or were not) translated into implementation.

### Data analysis

We conducted thematic analysis of the dataset. This comprised of 20 documents (Appendix 1), transcripts of 62 interviews (Appendix 2) and 17 observation sessions (Appendix 3). All these data pieces were brought together in Nvivo 12, which was also used to support analysis. We conducted open coding of the data. Codes with similar information were brought together to form themes.

The Multiple Streams Theory (MST) (11) and the Health Systems Strengthening (HSS) Framework (12) informed our initial coding and major themes. We refined the initial theory-informed coding and themes through repeated iterations of data analysis. Analysis iteratively informed our ongoing data collection. Using the underpinnings of MST, we explored the problems faced in SSP implementation and its beneficiaries during the COVID-19 pandemic. Under the policy stream, we explored the different policy options considered or selected by SSP in the pandemic response. The HSS Framework enabled us to explore how the different pillars of the health system (human resources, financing, delivery) and the organisational structures (funds’ flow, network hospitals, benefits package) of SSP facilitated or could have facilitated each other in the pandemic response.

### Reflexivity

Our personal experiences shape our views and interpretation of our observations. For example, one of the investigators (SAK) was infected with COVID-19 twice, six months apart. That might have influenced his views that the programme should have covered the pandemic. However, he did not need hospitalisation on either occasion. Also, SAK had previously worked at SSP and had a collegial rapporteur with the programme managers, putting him at risk of viewing the programme positively. SAK worked closely with AS and KC, who pointed out subjectivity in his work when noticed, and corrective measures were taken.

### Transferability

SSP is at a considerably advanced implementation stage compared to other SHP schemes launched by the provincial government of Gilgit Baltistan (GB) and the Federal Government of Pakistan (GOP) in other provinces. Since the GB and GOP programmes are scaling up their coverage and revising their benefits package, findings from this research might help them decide whether or not to cover COVID-19 or such health threats in their programmes’ design.

## Findings

Our findings are arranged under six themes. First, we describe the onset of the COVID-19 pandemic in Pakistan in the backdrop of the 2016 JEEP recommendations. Second, we present our findings on the overall government response. Third, we describe the health system’s response. Fourth, we present two types of SSP responses. The initial response (in 2020) was that COVID-19 was “not covered and not relevant to the programme”. In 2021, the response had changed to cover COVID-19. Finally, we describe stakeholders’ views on how the pandemic might influence the future trajectory of SSP.

### COVID-19 in the backdrop of JEEP recommendations

The COVID-19 pandemic caught Pakistan unprepared said a health systems expert. The system was “inept”, and many deficiencies were “exposed by the COVID-19 crises”. The major problems highlighted were inadequate hospital beds, ventilators and warehousing facilities:

> *“In COVID-19, an inadequate workforce…inadequate stocking of medicine and inadequate infection control measures were exposed”. [11: A health systems expert working at a development agency]*

Pakistan was caught off guard, said a senior official of Pakistan’s Ministry of National Health Services. The official reported that when the COVID-19 pandemic struck, they were working on three areas flagged in the JEEP by WHO, namely: (i) Integrated Disease Surveillance and Response (IDSR), (ii) antimicrobial resistance, and (iii) disease control at the entry points to the country:

> *“We were working on three areas…we were going in the right direction but caught midway”. [28: GoKP official]*

The prevention, detection and case management capacities were weak, according to a hospital manager. These weaknesses raised fears of catastrophic outcomes of the pandemic. There were no quarantine and isolation facilities, the health staff lacked basic personal protective equipment, and the limited ventilators in the province were already in use for critically ill patients:

> *“We had around about 600-700 ventilators throughout the province [for a population of 36 million]…The situation was scary. We did not even have the basic personal protective equipment for our COVID-19 isolation wards”. [42: Head of a public sector tertiary care hospital]*

Until the end of 2021, Pakistan reported 28,900 COVID-19-related deaths and a mortality rate of 6.6 per 100,000 population.(13) Deaths reported in KP were 5,930, with a mortality rate of 8 per 100,000 population.(13) The WHO considered these figures as grossly under-reported.(14) The estimated excess deaths for Pakistan and KP were 664,000 and 186,000, respectively.(14) The estimated excess mortality rates were 152.6 deaths per 100,000 population for Pakistan and 252.8 deaths per 100,000 population for KP.(14)

### COVID-19 and the overall government response

GOP led the COVID-19 response in consultation with the provincial governments. GOP had established the National Command and Operations Centre (NCOC), which articulated the entire response. A respondent in our research called NCOC, the “nerve centre” of Pakistan COVID-19 response.

A GOP respondent noted that at the onset of COVID-19, the [ex] Prime Minister of Pakistan [Imran Khan] announced a stimulus package of 1.3 trillion Pakistani Rupees (PKR). The money had allocation for social protection, health care and supporting businesses. The major chunk of the package provided relief to daily wage workers (PKR 200 billion) and low-income families (PKR 150 billion).(15,16)

A GOP official informed that the NCOC facilitated provincial health departments establishing COVID-19 quarantine centres and isolation wards in public hospitals. The cost of the COVID-19-related medical expenditures came from the fiscal stimulus announced by the [ex] Prime Minister of Pakistan [Imran Khan].

The documentary analysis (Pakistan Economic Survey 2020-21) showed that to address the shortage of medical equipment, GOP eliminated duties on the import of emergency medical equipment, established an emergency contingency fund (PKR 100 billion), and supported health and food supplies (PKR15 billion).(15,16) Additionally, the health sector obtained approvals of PKR 10.5 billion under the State Bank’s Refinancing Facility to keep hospitals [mostly private] afloat.(15,16)

### COVID-19 and the health system’s response

The estimated hospital bed capacity in Pakistan in 2017 was 109,132, with one bed per 1,580 people. In 2019, Pakistan had an estimated eight doctors per 10,000 population, compared to, for example, 12 in Iran and 35 in the UK.(17) Apart from human resources, it was acknowledged that the health system had other challenges, including deficiencies in health system data management and limited testing, tracing and quarantine services.

> *“Challenges were…deficiencies in the routine health system data and limited availability of the public health labs. There were limited facilities for testing, tracing, and quarantine*.*” [28: GoKP official]*

The government’s emphasis remained on public sector hospitals. A GoKP official shared two reasons for excluding the private sector from COVID-19 case management: (i) in public sector hospitals, where the government was in direct control of the response; and (ii) the private sector, which was kept as a reserve in case the public sector was overwhelmed.

> *“The government had established COVID wards in all the public sector hospitals*…*But people still had the option to visit a private hospital if they could afford it”. [45: Public sector hospital administrator]*

A private hospital’s manager however noted that not all private hospitals were allowed to treat COVID-19 patients. Except for some state-of-the-art private hospitals, the private sector was barred from treating COVID-19 patients. The private sector was kept as a reserve, said a GoKP official. However, the advocacy groups highlighted the government’s fear of a worsening pandemic and cost escalation at the loosely regulated private hospitals.

The public sector hospitals performed at full capacity. A hospital manager said their facility usually had a bed occupancy rate of 98%, which he noted was beyond the WHO recommended rate of 85-90%. They reportedly had taken several measures with limited resources to create a surge capacity for COVID-19 patients.

> *“We stopped our elective admissions as well as elective surgeries, in all the departments and got more beds available [for COVID-19 patients]” [42: Head of a public sector tertiary care hospital]*

Patients also avoided hospitals, creating capacity for COVID-19 patients, shared a public sector hospital’s manager:

> *“Due to the ongoing COVID, some patients are reluctant to get admitted. Otherwise, we have more than 80 per cent bed occupancy. [8: Manager at a public sector tertiary hospital]*

The pandemic had strained the budgets of the public sector hospitals. Hospitals had incurred liabilities on COVID-19 care, but a hospital manager noted that direct budgetary support from GoKP had decreased. They expected to raise revenue through SSP to bridge their budget gap:

> *“Due to the COVID-19 pandemic, we have created a financial liability of Rs*.*60 million. The direct government’s financial transfers are drying up, and we will have to generate revenue through the insurance programme [SSP]”. [7: A public sector tertiary care manager]*

But the SSP revenue was not forthcoming, as COVID-19 was not covered under the programme.

### COVID-19 and SSP’s initial response: not covered and not relevant

Throughout 2020, SSP did not offer COVID-19 treatment. When asked early in 2020 if SSP covered COVID-19, one officer responded, *“pandemics are not covered in insurance programmes”*. Another officer further said: “COVID-19 is not a part of this programme and hence not relevant”.

The insurer argued that epidemics and pandemics were the standard exclusion from insurance contracts and the GoKP officials agreed with their stance. Therefore, the role of SSP in the COVID-19 pandemic remained marginal (in 2020).

> *“Epidemics, endemics and pandemics cannot be covered in insurance schemes”. [2: SSP manager at GoKP]*

In contrast to the verbal response by SSP officials, the contract document between GoKP and the insurer did not mention epidemics and pandemics under the exclusions list. Another official of the GoKP noted that the discussions on coverage for COVID-19 under SSP at the beginning of the pandemic were inconclusive.

> *“There is no such effect of the corona crisis on this programme…it required additional strategising and funds which was not possible now”. [2: SSP manager at GoKP]*

And when asked if the insurance company would support COVID-19 treatment, a SLIC official noted that only non-COVID-19 admissions were allowed.

> *“Coverage for COVID-19 is not included in this programme, but we allowed non-COVID admissions”. [5: Manager at SLIC]*

However, there were no non-COVID-19 or routine admissions. All routine hospital admissions were stopped as a policy decision by GoKP. Therefore, the non-emergency SSP admissions had significantly decreased:

> *“Before the pandemic, there were 300-350 hospital admissions per day. During the pandemic, admissions reduced to 50 per day. This trend decreased the card utilisation”. [2: SSP manager at GoKP]*

Once routine services started, GoKP policy was to admit only COVID-19 negative patients. A SLIC official reported that the programme did not cover COVID-19 screening tests; hence patients had OOP expenditure. A SLIC manager noted the cost for [COVID-19] antibody and real-time reverse transcription polymerase chain reaction (RT-PCR) tests was PKR 1,000 and PKR 7,000, respectively.

> *“Due to the COVID-19 screening costs, the patients’ admission and surgery rate is very low. I think state life should contribute to such pandemic situations. In the seven thousand rupees of screening cost, state life should contribute three or four thousand”. [5: Manager at SLIC]*

### COVID-19 and SSP’s later response: to cover and stay relevant

As the pandemic progressed, the outlook of SSP changed, i.e., it started to finance COVID-19 treatment (from February 2021). A senior GoKP official presented two reasons for this change: (i) SSP utilisation rates had dropped significantly; and (ii) GoKP’s special fund for COVID-19 was depleting:

> *“The government asked us to identify the top 10 private hospitals to provide services to COVID-19 patients under SSP”. [1: a senior GoKP official]*

It marked a departure from the earlier stance of the programme that “COVID-19 was not covered and not relevant”. A GoKP official informed that in 2021, SSP had 1,002 COVID-19-related admissions, which were 0.2 % of the total hospital admissions (N=544,841). However, the official informed that the average cost for a COVID-19 patient was more than thrice the average cost of a non-COVID-19 admission.

A GoKP official noted that the COVID-19 coverage under SSP, once started, was unlimited, i.e., there was no upper limit on expenditure. SLIC officials still maintained that pandemics were not an insurable risk, and the COVID-19-related hospital bills were paid from the reserve fund. A GoKP official informed that the reserve fund was established for high-cost procedures like kidney and liver transplants and provided additional coverage for critically ill patients whose standard coverage had expired.

Hence to cover COVID-19, the benefits package or the premium were not revised. As per the GoKP and SLIC contract, the insurer’s profit depended on savings of the primary pool (premium). The company worked as a third-party fund administrator for the reserve fund. SLIC did not assume any profit or loss for the reserve fund.

Though the COVID-19 coverage did not accrue any cost to the insurer, their representative did not sound positive. This may have been due to a potential increase in their workload or the absence of a financial incentive for the company:

> *“With the 100% coverage, now they are in haste in adding more and more services…they have also included COVID-19 coverage from the reserve fund…you cannot just replace all the parallel programmes with the insurance scheme”. [6: Manager at SLIC]*

In contrast with the SSP coverage, a SLIC official reported that they had extended coverage for COVID-19 to their corporate clients. Corporate clients were public or private sector employers who had purchased group life insurance from SLIC for their staff, and their health coverages linked to their unit-linked insurance plans.

SSP did not play a role in the COVID-19 vaccination either. The COVID-19 vaccination formally started on 3 February 2021 with a donation of 1.5 million Sinopharm doses from China.(170) The global shortage of the vaccine was cited as a reason by a GoKP official for delays in the vaccination rollout:

> *“Pakistan started vaccination in February 2021. We started vaccination on a priority basis for 60 plus population and those working in health care. As vaccines became available, younger age groups were vaccinated”. [A senior GoKP official]*

The GOP and GoKP used the public sector infrastructure for the COVID-19 vaccination rollout. As more vaccines became available, GoKP considered engaging the private sector. A SLIC official informed that GoKP wished to engage the private hospitals under the SSP network in the vaccination rollout. Subsequently, SLIC wrote to the private hospitals to arrange for administering COVID-19 vaccines. GoKP would provide the vaccines, and the hospitals would administer them free of cost.

> *“Now another intervention they introduced is involving the programme in the COVID-19 vaccination. This is a big problem. They are putting all the eggs in one basket”. [6: Manager at SLIC]*

The insurance officials considered the private sector’s response to the vaccine proposal positive. Still, the government did not follow through with those plans, and the reasons were not known to the SLIC manager.

### SSP beyond COVID-19

The preceding sections reveal that COVID-19 changed policymakers’ and managers’ understanding of SSP’s role. The pandemic also re-emphasised the role of international partnerships in developing resilient health systems and responding to global health threats. A manager of the German Development Bank (KfW) suggested the pandemic might change the working relationship between GoKP and its international partners, especially KfW. The manager highlighted that health was dropped from the [Pak-German] cooperation priority list, at the request of the Pakistani side, when Nawaz Shareef [the then Prime Minister of Pakistan] met with [then] Chancellor Merkel. Highlighting the small percentage of GDP allocated to health, the KfW manager opined Pakistan could not handle disasters like the COVID-19 pandemic alone and needed external support.

> *“The health sector, for the time being, is not a priority sector anymore in Pakistan…because of the COVID pandemic…chances are that health might be re-introduced as a priority sector in Pakistan. [16: A KfW Manager]*

KfW managers highlighted that the German Government had allocated some money to assess the pandemic preparedness of Pakistan, and results from that study might change the nature of the partnership. Moreover, the pandemic also influenced the current engagement of KfW with the programme. For example, the KfW consultants working on designing the outpatient department (OPD) coverage under SSP noted that the COVID-19 pandemic might change their design, which they initially thought would cover chronic non-communicable diseases only.

> *“Launching an OPD component during a pandemic without covering COVID-19 or other infectious diseases would be hard to justify”*.*[29: A member of the KfW consulting team for the OPD project]*

An official of SSP suggested that the programme covered 100% of the province population, and the utilisation trends in its data could be used to pick epidemics and outbreaks. He suggested that in the long run, SSP could serve as the nerve centre for future pandemics, as was the NCOC in COVID-19. A health system specialist working with a development organisation did not share this optimism however because, according to him, the programme lacked the technical capacity to analyse and interpret the insurance utilisation data [for surveillance].

## Discussion

### Summary of the key findings

COVID-19 exposed the protection gaps in SSP coverage at several stages as the pandemic progressed, namely: (1) SSP refused to cover COVID-19 treatment’ (2) the ban on routine medical care complicated the continuity of care for many SSP patients; (3) once routine services restarted, SSP patients had to pay for COVID-19 testing, as SSP did not cover it; (4) when the pandemic entered year 3, SSP started to cover COVID-19 treatment; (5) the reserve fund earmarked for other life-saving procedures was redirected to COVID-19 treatment; and (6) the inconsistencies in the programme’s pandemic response called for rethinking and reevaluation of the protection claims and highlighted the need for better pandemic financing strategies.

### Strengths and limitations

Our enquiry regarding the initiation and implementation of SSP was planned when the COVID-19 pandemic started. The qualitative nature of our work enabled us to add questions related the programme’s role in the COVID-19 response. However, we could not cover the full range of GoKP’s COVID-19 response. That was beyond the ambit of our enquiry. Although we had interviews representatives of patient advocacy groups, including a cancer support group and a disabled persons association, we did not have the perspective of any COVID-19 patients. Apart from public health concerns, our study design and ethics approvals did not include direct interaction with patients.

### Interpretation in view of the broader literature

The COVID-19 pandemic tested the resilience of health systems and social protection regimes across the globe.(18–20) In some places, it showed the advantages of having Universal Health Coverage (UHC); in others, it intensified the calls for UHC, elsewhere; it reversed decades of delicate work toward UHC.(18) Additionally, it further authenticated the notion that UHC and GHS were inseparable and intertwined.(16)

GHS refers to the proactive and reactive activities to minimise vulnerability to acute public health events that endanger the collective health of populations living across geographical regions and international boundaries.(20) Countries with UHC and better GHS preparedness managed the pandemic well. Countries missing either UHC or GHS did not fair well. Others with limited UHC and GHS suffered the most.(19)

Pakistan’s GHS Index 2021 score was 30.4 [the score range is 0-100 with 100 being ideal], and the country was ranked 130 among 195 countries. The country was ranked 192 for its response capacities and 150 under risk subcategories.(2,5) Our study supports the GHS and JEEP findings that the country lacked plans for private-sector engagements in responding to health threats.(5) Pakistan did not have UHC either.(21) Lack of reporting systems might be one reasons that the incidence and mortality rates in Pakistan were grossly under-reported.(14)

Though an official described that COVID-19 was “not covered and not relevant”, the very nature of the crises and the programme made it relevant.(22,23) SHP had a significant role at the beginning of the pandemic (2020) when the economy was in a tailspin, inflation rose, and around 13 million jobs were lost.(15,16) SHP means guaranteed access for people to address their health needs irrespective of their ability to pay.(24) Else, health care expenditure in the face of financial downturn and lost incomes would push people into poverty.(24)

Various social protection interventions were undertaken under Ehsas Programme, including: (i) Ehsas emergency cash transfer; (ii) Ehsas Nash-O-Numa (nutrition support programme);and (iii) Ehsas Langar (free food kitchens).(16) However, the role of an important component, i.e. SHP, was not as evident as the Ehsas Programme. Since SSP refused services to COVID-19 patients in the acute phase of the pandemic (2020), it remained a remote player in the pandemic response.

It can be argued that SSP did not add to the health system’s resilience. Resilience refers to the ability of a system, community or society exposed to hazards to resist, absorb, accommodate, adapt to, transform and recover from the effects of a hazard in a timely and efficient manner, including through the preservation and restoration of its essential basic structures and functions through risk management.(20) The role of SSP was in stark contrast with similar public-funded health insurance programmes.(20,25,26)

For example, in the Philippines, PhilHealth offered services to COVID-19 patients.(25) Initially, PhilHealth covered people under the existing benefits package, but as the epidemiology and case management guidelines of COVID-19 became clearer, PhilHealth introduced a dedicated COVID-19 package.(25) Both suspected and confirmed cases were covered with defined tariffs.(25) The Indian public-funded Pradhan Mantri Jan Arogya Yojana (PMJAY) also covered treatment for COVID-19 at both public and private sector hospitals.(26) By comparison to PhilHealth and PMJAY, SSP did not do well.

The Social Health Insurance (SHI) programmes in Europe went much further than SSP, PhilHealth or PMJAY.(27) The SHI programmes in different European countries covered COVID-19-related tests, ambulatory services, inpatient care and vaccines. These programmes also added innovative and agile solutions.(27) The SHI funds paid for teleconsultations, participated in contact tracing, established new access pathways and offered compensations to participating physicians who lost income due to disruption of routine services.(27) Additionally, the SHI funds adopted different mechanisms to maintain the fund’s solvency yet ensured free access to health care.(27) Considering these differences, SSP has a long way to get established as a true SHP or a SHI programme.

#### Implications for policy, practice and research

Our work has indirectly demonstrated that the pandemic affected access to health care and that SSP beneficiaries were exposed to OOP payments. We could not ascertain whether the OOP expenditure made people forgo treatment. Due to the qualitative nature of work, we could not answer several important questions, including: (1) how did the SSP services utilisation compare during the pandemic to the pre-pandemic numbers; (2) how did refusing services to COVID-19 affect the balance sheet of the insurance company; and (3) did the diversion of the SSP Reserve Fund towards COVID-19 patients affect other patients? We recommend that these questions should be considered for future research

Implications for policy and practice of SSP and other SHP programmes’ policymakers consideration were: (I) not the name, but the policies and their pro-patient interpretation make programmes SHP or otherwise, (II) while SSP in KP is considering coverage for outpatient services under a fund management modality, it is worth remembering that insurance firms might be more cost-savvy when working on a premium than fixed admin costs, (III) as KfW develops the outpatient component of SSP, it is essential to take the JEEP recommendations on board, and (IV) considering insurer’s incentive to minimise losses and the government’s precarious position to respond to a situation like pandemics, newer financing modalities like catastrophic bonds, or parametric insurance instruments should be developed. Further research is recommended to operationalise these recommendations, bridge the protection gaps and improve the SSP response to future health threats.

## Conclusion

GoKP started to utilise the SSP network hospitals more than a year after COVID-19 had arrived in Pakistan. The initial refusal to cover COVID-19 and later coverage through the SSP Reserve Fund raised several questions.

Initial refusal of SSP to cover COVID-19 treatment and its underlying assumptions did not present a congruent rationale for two reasons. First, insurer justified the refusal of COVID-19 treatment, arguing that pandemics were a standard exclusion from insurance contracts. The question was whether the SSP contract was commercial or socially driven. SSP officials sided with the insurer’s interpretation, but the description of SHP did not suit this commercial interpretation. Second, if pandemics were a standard exclusion, how could the insurance industry, including SLIC, provide COVID-19 treatment to corporate clients? Refusing services to the vulnerable in a pandemic under a publicly funded social protection programme might not promote equity and social protection.

The average cost of treatment for COVID-19 was much higher than the average cost of non-COVID cases under SSP. Many patients who had COVID-19 treatment before SSP started to pay for it might have experienced OOP and catastrophic health expenditures. On the one hand, the COVID-19-related treatment might be expensive, but on the other hand, the insurer might not have acted vigorously to cut costs, as they had no incentive to save the SSP Reserve Fund money. Using the SSP Reserve Fund insulated the primary insurance pool (money paid as premium), which apparently kept the insurer profitable.

## Data Availability

The datasets used and analysed during the current study are available from the corresponding author on reasonable request.

## Declarations

### Ethics approval and consent to participate

We had ethics approval from the Ethics Committee at the University of Edinburgh and local ethics approval from the Khyber Medical University. We have complied with the ethics regulation of the approval bodies at all stages of the research and have maintained a written record of informed consent for the study participants.

### Consent for publication

Our work does not include any potential identifiers of our participants, and we had the consent of the participants to publish our work based on their participation. We did not use any copyrighted material in this paper.

### Competing interests

We have no competing interests.

### Funding

The work was made possible through a PhD funding from the Higher Education Commission of Pakistan. The funder did not influence the study design, data collection, analysis or reporting.

### Authors’ contributions

SAK developed the research plan and led the data collection, analysis and write-up. KC and AS contributed to refining the study design advised on the data collection, analysis, and interpretation, and helped refine the manuscript. All authors read and approved the final document.

## Acknowledgements

We acknowledge the SSP Director for supporting our work.

## Appendix 1: List of documents included in the documentary analysis

### SSP policy and implementation documents

1. Planning Commission Form 1 for SSP.
2. Request for proposal (RFP) document, for hiring insurance company for 100% population coverage [Phase 4].
3. GoKP and SLIC contracts for SSP.
4. Khyber Pakhtunkhwa Universal Health Coverage [Draf] Act, 2022.
5. KfW commissioned feasibility study for SSP (KfW Phase 1).
6. Inception report for the OPD component of SSP (KfW Phase 2).
7. Hiring of consultant for feasibility study for social health protection project Phase III (digitilisation of SSP: KfW Phase 3).
8. Guiding document for the development of a roadmap towards achieving universal coverage.
9. List of hospitals working with SSP.
10. Joint review of the SSP.
11. Baseline for communication strategy of Sehat Card Plus-KP.
12. First-Year Report on services’ utilisation under the universal population coverage conferred by the social health protection initiative (Sehat Card Plus) in Khyber Pakhtunkhwa, Pakistan.

### Broader policy documents

13 Pakistan Economic Survey 2020-21.
14 National Health Accounts 2017-18.
15 Health Policy Khyber Pakhtunkhwa.
16 Pakistan National Health Vision, 2025.
17 Government of Khyber Pakhtunkhwa: White Paper; Fiscal Year 2021-22.
18 Moving together to build a healthier Pakistan: Khyber Pakhtunkhwa. In: Sensitisation and Situation Analysis Workshop: provincial localisation of UHC Benefit Package.
19 Burden of Disease, Khyber Pakhtunkhwa. In: Sensitisation and Situation Analysis Workshop: provincial localisation of UHC Benefit Package.
20 UHC Benefit Package of Pakistan: Essential package of health services at community and primary healthcare centre level based on Disease Control Priorities-Edition 3.

## Appendix 2: Distribution of in-depth interviews at policy and implementation level stakeholders of SSP

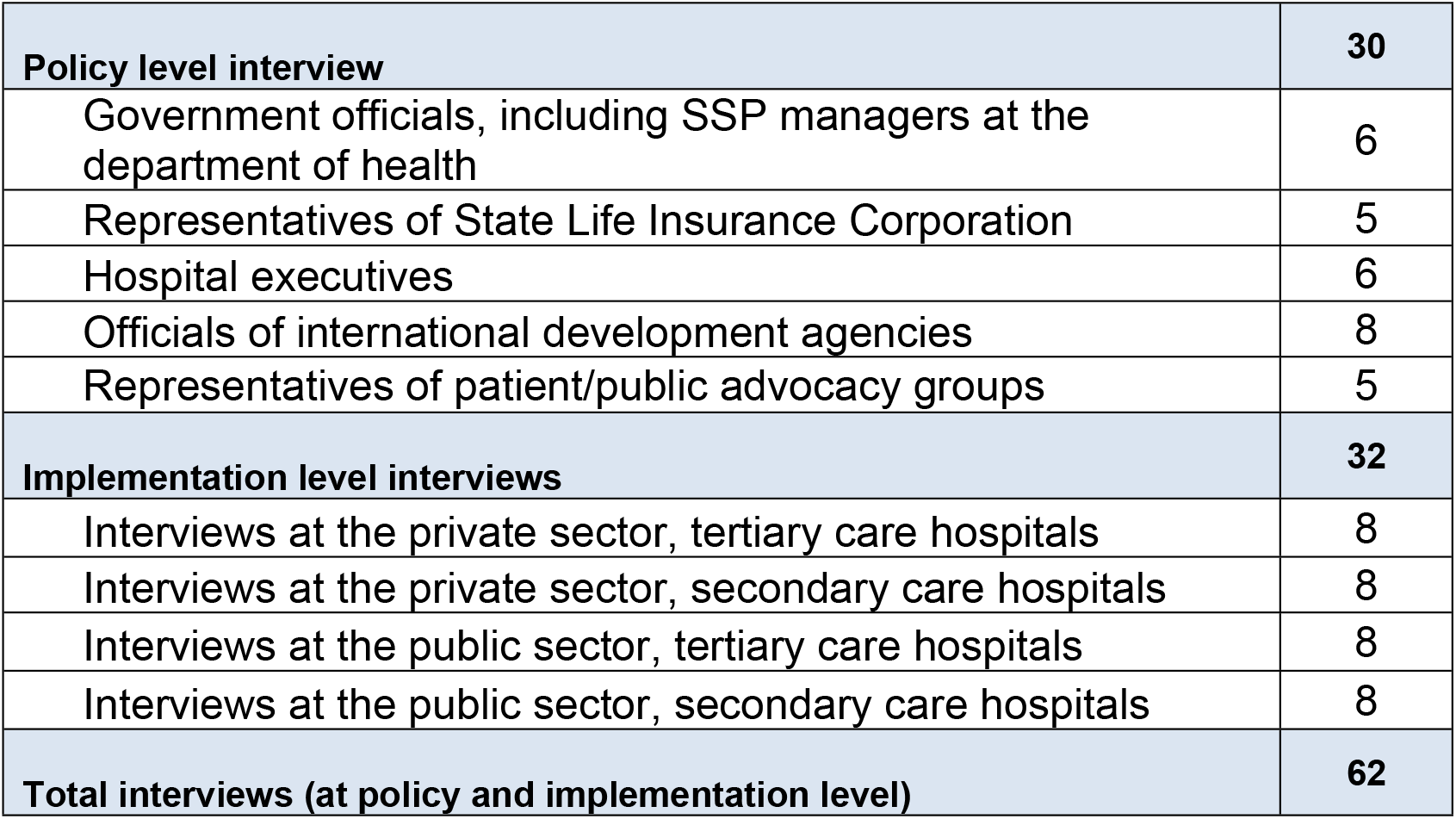

## Appendix 3: Distribution of non-participant observations at SSP policy meetings and implementation sites

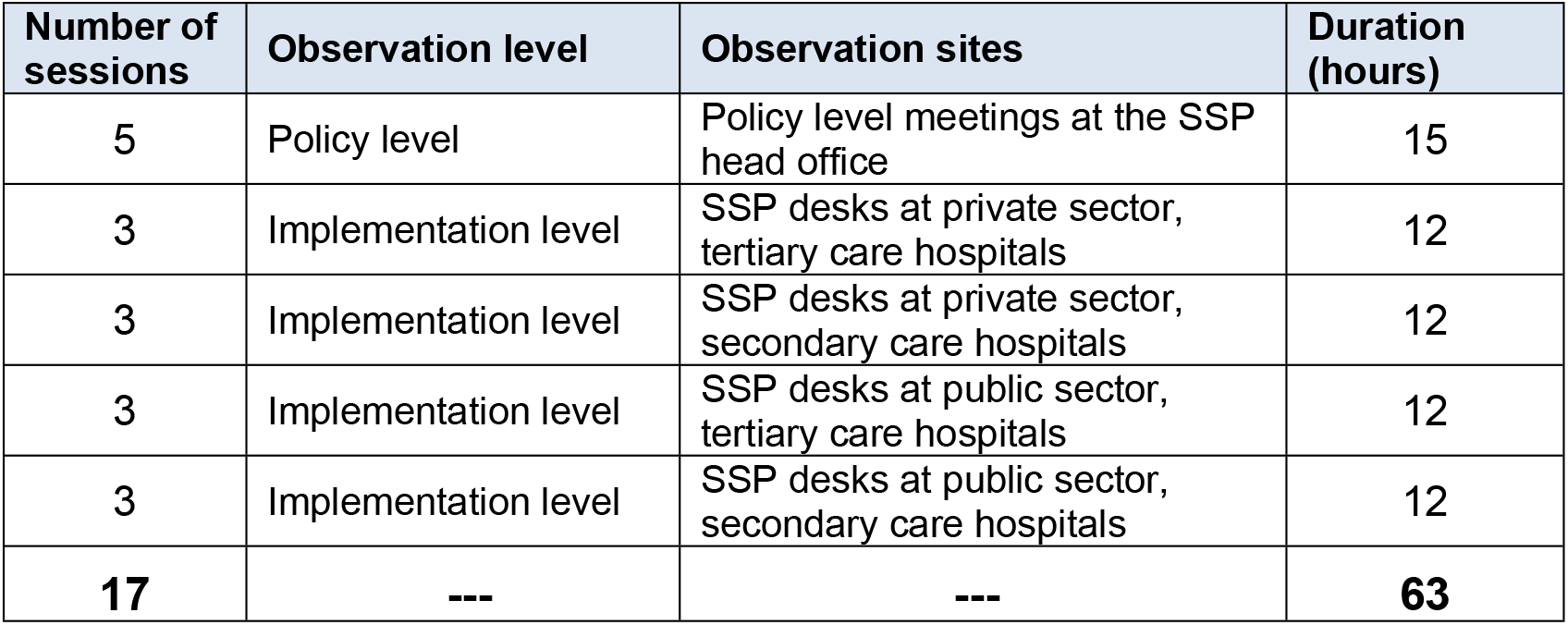

**Appendix 3: Distribution of non-participant observations at SSP policy meetings and implementation sites.**
| **Number of sessions** | **Observation level** | **Observation sites** | **Duration (hours)** |
| --- | --- | --- | --- |
| 5 | Policy level | Policy level meetings at the SSP head office | 15 |
| 3 | Implementation level | SSP desks at private sector, tertiary care hospitals | 12 |
| 3 | Implementation level | SSP desks at private sector, secondary care hospitals | 12 |
| 3 | Implementation level | SSP desks at public sector, tertiary care hospitals | 12 |
| 3 | Implementation level | SSP desks at public sector, secondary care hospitals | 12 |
| **17** | **---** | **---** | **63** |

## Notes

### Competing Interest Statement

The authors have declared no competing interest.

### Author Declarations

We had ethics approval from the Ethics Committee at the University of Edinburgh and local ethics approval from the Khyber Medical University. We complied with the ethics regulations of informed consent, participants’ autonomy, confidentiality, and right to withdraw.

